# Cardiorespiratory fitness and cerebral blood flow in cognitively normal older adults and individuals with coronary artery disease: the AGUEDA and Heart-Brain projects

**DOI:** 10.64898/2026.03.03.26347189

**Authors:** Lucía Sánchez-Aranda, Kai de Geus, Patricio Solis-Urra, Javier Sanchez-Martinez, Angel Toval, Isabel Martín-Fuentes, Javier Fernández-Ortega, Rosa María Alonso-Cuenca, Beatriz Fernandez-Gamez, Marcos Olvera-Rojas, Andrea Coca-Pulido, Anna Carlén, Eduardo Moreno-Escobar, Rocío García-Orta, Kay Jann, Kirk Erickson, Irene Esteban-Cornejo, Francisco B Ortega

**Author notes:** These authors contributed equally to this work (Lucía Sánchez-Aranda, Kai de Geus). Joint senior authors (Irene Esteban-Cornejo, Francisco B Ortega). Correspondence: Lucía Sánchez-Aranda, Irene Esteban-Cornejo and Francisco B. Ortega. E-mail address, and, Full address: Department of Physical and Sports Education, Faculty of Sport Sciences, University of Granada, Granada, Spain.

## Abstract

Age and coronary artery disease (CAD) are known risk factors for cognitive decline and dementia, in which cerebral blood flow (CBF) has as a key role. Cardiorespiratory fitness (CRF) has shown consistent links with brain health and dementia, though its association with CBF and whether it differs depending on age or disease status remains limited. The main aim of this study was to examine the association of CRF, assessed through the six-minute walk test (6MWT) and peak oxygen uptake (VO₂peak), with CBF in cognitively normal older adults and individuals with CAD. We hypothesized that CRF will be positively associated with global and regional CBF.

Seventy-nine cognitively normal older adults from the AGUEDA trial and 84 individuals with CAD from the Heart-Brain trial were included in this cross-sectional analysis. Participants underwent 6MWT, and a 3D arterial spin labelling magnetic resonance imaging scan to assess global and regional CBF. In the Heart-Brain project, participants additionally conducted a cardiopulmonary exercise test from which VO_2_peak was determined.

In the Heart-Brain sample, after adjusting for age, sex, education, mean arterial pressure and APOE4, CRF was positively associated with global CBF (6MWT: β=0.26, P=0.04; VO_2_peak: β=0.33, P=0.02). At a regional level, CRF was positively associated with CBF in the posterior cingulate cortex (6MWT: β=0.26, P=0.04; VO_2_peak: β=0.31, P=0.02), the anterior cingulate cortex (6MWT: β=0.29, P=0.02), the precuneus (6MWT: β=0.28, P=0.02; VO_2_peak: β=0.32, P=0.01) and the hippocampus (VO_2_peak: β=0.29, P=0.03). No significant associations were observed in the AGUEDA sample (all P>0.05). When adding body mass index (BMI) to the models, the associations were no longer statistically significant in either sample. The association between VO_2_peak and CBF was significantly mediated (i.e. indirect effect) by BMI (Indirect effect: β=0.250 (95% CI 0.02;0.486), percentage of mediation=72.67%).

CRF was positively associated with CBF in individuals with CAD, but not in cognitively normal older adults. Interestingly, the association of CRF with CBF was largely explained and mediated by BMI. Further studies are warranted to clarify the role of CRF and BMI in relation to CBF, the mechanisms involved and the implications for dementia risk prevention in older adults and individuals with CAD.

## Introduction

As the proportion of the aged population increases, with more than one billion individuals being 60 years or older worldwide, the prevalence of age-related health conditions, including dementia, also rises.^1^ In addition to age, numerous factors contribute to the onset and progression of dementia.^2^ Cardiovascular disease (CVD) is strongly linked to an increased risk of cognitive decline^3,4^ and dementia,^5–7^ and this association is especially evident in coronary artery disease (CAD),^8,9^ the most common form of CVD.^10^

Decreases in cerebral blood flow (CBF - hereafter grey matter only, unless otherwise indicated), which entails a reduced supply of oxygen and nutrients to brain tissue,^11,12^ have been associated with an increased risk of dementia in the general population.^13^ CBF has been shown to decrease with age,^14–16^ with more exacerbated reductions in individuals with CVD,^17–20^ making it a plausible candidate to explain the interplay between the heart and the brain. Indeed, well-known cardiovascular risk factors are linked to more probable development of dementia,^21–23^ and, particularly, high body mass index (BMI), has been consistently associated with lower CBF.^24,25^

In this context, cardiorespiratory fitness (CRF) emerges as a potential protective factor within the heart-brain axis. CRF is not only a well-known indicator of overall cardiovascular health and a strong predictor of all-cause mortality,^26–28^ but greater CRF is also related to lower risk of cognitive decline and dementia.^29,30^ Recent studies have shown CRF to be positively associated with cognitive function^31,32^ and brain structure^33,34^ in general older adult population. Moreover, we have observed larger hippocampal grey matter volume in individuals with CAD that have higher CRF levels.^35^ However, whether a link exists between CRF and CBF remains unclear. In contrast to the negative association reported by Olivo et al.,^36^ two studies in middle aged and older adults^37,38^ and one in individuals with CAD,^39^ found a positive association between CRF and CBF, whereas two others found no significant relationship in general aging samples.^40,41^

The aim of the present study was to assess the association of CRF, as measured with the 6 minute-walk test (6MWT) and peak oxygen uptake (VO_2_peak), with CBF (global and in the hippocampus, the precuneus, the posterior cingulate and the anterior cingulate cortex) in a sample of cognitively normal older adults and a sample of individuals with CAD. We hypothesized that, higher CRF would be positively associated with CBF (at both, global and regional levels), in cognitively normal older adults and individuals with CAD. Furthermore, we hypothesized that the association would be stronger in the sample of individuals with CAD, given their compromised cardiovascular function and increased burden of brain pathology, in which CRF could play a major role.

## Materials and methods

### Study design and participants

Baseline data from two randomized controlled trials (RCT) were used for this study. Ninety-one cognitively healthy adults, aged 65-80 years old, participated in the baseline assessments of the “Active Gains in brain Using Exercise During Aging” (AGUEDA) RCT (ClinicalTrials.gov, Identifier: NCT05186090; Submission date: December 22, 2021), and 105 individuals with CAD, aged 50-75 years, participated in the Heart-Brain RCT (ClinicalTrials.gov, identifier: NCT06214624; Submission date: December 22nd, 2023), with no participant overlapping between trials. From these, a total of 10 (one from AGUEDA RCT and nine from Heart-Brain RCT) had no magnetic resonance imaging (MRI) scan, 23 had no valid ASL sequence (11 from AGUEDA RCT and 12 from Heart-Brain RCT), resulting in a final sample of a 163 participants for this cross-sectional analysis (AGUEDA = 79, Heart-Brain = 84).

Both trials were in accordance with the principles of the Declaration of Helsinki and have been approved by the Research Ethics Board of the Andalusian Health Service (CEIM/CEI Provincial de Granada; AGUEDA: #2317-N-19 and Heart-Brain: #1776-N-21). All participants gave written informed consent. Detailed information about AGUEDA and Heart-Brain RCTs can be found at Solis-Urra et al.^42^ and Toval et al.,^43^ respectively.

### Inclusion/exclusion criteria

Inclusion criteria have been previously defined in detail in the protocol articles.^42,43^ In brief, inclusion and exclusion criteria for each project were as followed:

Participants from the AGUEDA RCT were defined as (i) cognitively normal according to the Spanish version of the modified Telephone Interview for Cognitive Status (score ≥26), the Montreal Cognitive Assessment, and the Mini Mental State Examination and (ii) physically inactive (i.e. not participating in any resistance exercise programs in the last six months and accumulating < 600 METs/week of moderate-vigorous physical activity) as assessed by self-reported questions about structured exercise and physical activity. All participants were medically cleared by a sports medicine physician, ensuring that they had no physical impairments and psychiatric or neurological conditions.

Participants from the Heart-Brain RCT were defined as (i) individuals with CAD (phase III) with a stable medical treatment, aged between 50-75 years; (ii) with preserved left ventricular ejection fraction (≥45%); (iii) with no major cognitive impairment according to the Spanish version of the modified Telephone Interview for Cognitive Status (score ≥26) and (iv) physically inactive (not meeting the WHO recommendations, thus not participating in a planned and structured exercise program at least three days per week for the last three months) as assessed by self-reported questions about structured exercise and physical activity.

### Cardiorespiratory fitness

Cardiopulmonary exercise testing (CPET) with gas exchange analysis is considered the gold standard for assessing CRF via peak oxygen uptake (VO₂peak). However, CPET is often expensive and time-consuming, what limits it usage in large scale studies. Besides, older adults may find difficulties to achieve maximal performance, potentially limiting the accuracy of the data. In contrast, the six-minute walk test (6MWT) is as a reliable, low-cost, and time-efficient alternative for estimating CRF in clinical and aging populations. Notably, previous research has shown that performance on the 6MWT correlates well with VO₂peak,^44^ making it a suitable proxy in contexts where CPET is not feasible.

We used the 6MWT in both samples as sub-maximal indicator of CRF, and VO_2_peak in the CAD sample, as gold-standard physiological measure of CRF.

#### Six-minute walk test

CRF was assessed using the 6MWT,^45^ a well-established tool for evaluating CRF,^44^ in both samples. Participants were instructed to walk as much as possible during six minutes, along a 4.57 × 15-meter flat rectangle. Standardized instructions and verbal encouragement were provided throughout the test. The total distance walked was measured in meters, with larger distance indicating better performance.

#### Peak oxygen uptake

Participants from the Heart-Brain trial performed a standardized incremental cardiopulmonary exercise test (CPET) on a treadmill (h/p/cosmos, Nussdorf, Germany), following the American College of Sports Medicine (ACSM) guidelines,^46^ with registration of respiratory gases (Omnical, Maastricht Instruments, Maastricht, the Netherlands; time resolution: five seconds). The protocol included a three-minute warm-up with progressively increasing speed until 4.8 km/h followed by 1% increase of the inclination every 30 seconds until volitional exhaustion or other indication for terminating the test. Peak oxygen uptake (VO_2_peak; mL/min) was calculated as the highest rolling 30-second VO_2_ average during exercise, and was then normalized to the individuals’ body mass to get VO_2_peak in mL/kg/min. Further details on CPET processing can be found in Carlén et al.^35^

### MRI acquisition and processing

MRI sequences, processing, and visual inspection and quality control were equally performed for both projects.

A 3.0 Tesla Siemens Magnetom Prisma MRI scanner (Siemens Medical Solutions, Erlangen, Germany) equipped with a 64-channel phased-array head coil was used for image acquisition. A modified background-suppressed four-delay pseudo–Continuous Arterial Spin Labeling (pCASL) sequence based on the protocol by Wang et al.,^47^ including proton density calibration image (M0) and 9 label-control pairs, was performed to obtain CBF measurements. Participants were advised to remain awake during the scan. Sequence parameters were: labeling duration = 1500 ms, post-labeling delays: 2×500, 2×1000, 2×1500, 3×2000 ms, TR = 4300ms, TE = 23.64ms, flip angle = 120, FOV = 300 mm x 300 mm, slice thickness = 2.5 mm, voxel resolution = 3.1 × 3.1 × 2.5 mm. A sagittal 3D T1 weighted image (TR = 2400, TE = 2.31, number of slices = 224, voxel resolution = 0.8 x 0.8 x 0.8 mm) was acquired and processed for registration and segmentation purposes.

First, T1 weighted scans were preprocessed using FSL’s ANAT pipeline.^48^ Then, two stepwise coregistration was performed to align the T1 weighted image and Arterial Spin Labelling (ASL) M0. Mri_coreg from Freesurfer (version 7.4.1)^49^ performed a linear registration between the two volumes. The resultant transformation matrix was then used as initial transformation matrix for running FSL’s FLIRT^50–52^ boundary-based coregistration, with six degrees of freedom rigid body transformation.

Finally, ASL to structural transformation matrix coming from FSL’s FLIRT was entered into oxford_asl command, together with ASL data and FSL’s ANAT outputs to get CBF maps. Oxford_asl uses the FSL’s FABBER ASL package and Bayesian Inference to invert the kinetic model for ASL MRI (BASIL).^53,54^ Previously described sequence parameters (post-labeling delays, TR and TE) and labelling efficiency (set at 0.7) were specified as input to the kinetic models for estimating CBF. Additional input parameters were kept with default settings corresponding to pCASL acquisition. Partial volume correction was used to report global CBF.^55^ All CBF values refer to grey matter only.

Region specific analysis was performed using Freesurfer’s outcomes from recon-all longitudinal processing, which has been described elsewhere.^56^ First, FreeSurfer’s brain image was coregistered to the ASL image using epi_reg.^52^ The resulting transformation matrix was then applied to the aparc+aseg segmentation image. Then, cortical and subcortical grey matter regions were extracted with fslmaths command, and the mean CBF for each region of interest (ROI) was calculated separately for each hemisphere by averaging CBF values of all voxels within the ROI using the fslstats command. Due to high correlation between hemispheres (r Pearson correlation values between 0.8 and 0.9, p<0.001) for the regions included in this study (hippocampus, posterior cingulate cortex, anterior cingulate cortex and precuneus), values from both hemispheres were later averaged. These regions were selected given their stablished relationship with cognition and dementia development.^57–61^

Mean native space difference maps (i.e. control minus label) for each participant were visually inspected,^62,63^ by two independent raters (LSA and KdG), to ensure data quality. Particular attention was paid to ensure that there were no: 1) excessive motion resulting in spurious effects in difference map; 2) maps with brain territories which did not appear to be perfused; 3) maps with perfusion difference between hemispheres; and 4) faulty scans due to wrong placement of the ASL imaging slab. Further quality control procedures included the calculation of grey matter CBF spatial coefficient of variation (CoV), as previously described by Mutsaerts et al.^64^ grey matter CBF maps were ranked into 3 levels according to their grey matter CBF CoV: ‘good’ (CoV < 0.6, CBF signal predominates artefacts); (ii) ‘acceptable’ (0.6 ≤ CoV ≤ 0.8, both CBF signal and artefacts visible); or (iii) ‘bad’ (CoV > 0.8, artefacts predominate CBF signal). CBF maps were later reviewed, and CoV-based categorization of the images were corrected if necessary. Mean native space difference maps that met at least one of the previously defined criteria, and CBF maps ranked with CoV acceptable or bad, were excluded from the analyses. Further details on the number of scans excluded and the reasons for ineligibility are provided in Supplementary Table S1.

### Covariates

Based on prior literature, we included the following covariates: age,^14–16^ sex,^14,16^ level of education (primary, secondary, university),^65–67^ body mass index (BMI; kg/m²),^24^ mean arterial blood pressure (MAP; mmHg),^68^ and apolipoprotein E ε4 allele (APOE4) carrier status (carriers vs. non-carriers).^69,70^

Sex was categorized as male or female based on self-reported information. Level of education was registered as the self-reported highest degree of education reached and categorized as: primary school (1), secondary school (2) or university (3). BMI was calculated as the participant’s weight divided by the participant’s square height, both measured using a SECA 225 (Seca, Hamburg, Germany) stadiometer and scale. Blood pressure was measured at rest in a sitting position with an automatic sphygmomanometer. For MAP calculation we summed two times diastolic blood pressure plus systolic blood pressure and then divided by three. The genotyping of APOE4 for the AGUEDA study was carried out at GENYO, Granada (Spain) using KASPar® assays (LGC Genomics, Hoddesdon, UK) according to the manufacturer’s protocol for SNP analysis,^71^ ensuring standardized protocols to ensure the quality and accuracy of the results. For the Heart-Brain sample, genotypes for the two APOE4 single nucleotide polymorphisms (rs429358 and rs7412), were performed on blood samples using TaqMan genotyping assays.^72^ Participants with at least one ε4 allele (ε2/ε4, ε3/ε4 or ε4/ε4) were classified as APOE4 carriers.

### Statistical analysis

All statistical analyses were performed using R studio version 4.5.1. Visual inspection of histograms and Kolmogorov-Smirnov test (P>0.05) were performed to determine the normal distribution of the variables. Pearson correlations were calculated for each sample to test the associations between CRF, CBF and the covariates included in the study.

Multiple linear regressions were fitted to test the associations between CRF (6MWT in both samples and VO_2_peak in the Heart-Brain sample) and CBF, analysed in each sample separately. First model included age, sex, education, APOE4 carrier status and MAP as covariates, whereas second model was additionally adjusted for BMI.

Further exploratory analyses included the mediation effect of BMI in the association between CRF and CBF, using Structural Equation Model (SEM) through lavaan package.^73^ BMI was introduced as potential mediator on the association between CBF and CRF. We included age, sex, education, APOE4 carrier status and MAP in the model.

## Results

### Descriptive Characteristics

The descriptive characteristics of the study sample are shown in Table 1. Briefly, AGUEDA sample had a mean age of 71.4±3.9 years whereas Heart-Brain sample had a mean age of 62.4±6.8. AGUEDA sample had a higher proportion of women than Heart-Brain sample (56*%* and 23*%*, respectively), and lower 6MWT performance.

**Table 1.** Descriptive characteristics of the study samples.

| Characteristic | AGUEDA<br>N = 79 | Heart-Brain<br>N = 84 | P value |
| --- | --- | --- | --- |
| Sex, n (%) |  |  | **<0.001** |
| Male | 35 (44%) | 65 (77%) |  |
| Female | 44 (56%) | 19 (23%) |  |
| Age (years) | 71.44 ± 3.97 | 62.36 ± 6.81 | **<0.001** |
| Education level, n (%) |  |  | 0.5 |
| Primary studies | 13 (16%) | 17 (20%) |  |
| Secondary studies | 28 (35%) | 34 (40%) |  |
| University studies | 38 (48%) | 33 (39%) |  |
| Apolipoprotein E-ε4 carriership, n (%) |  |  | 0.5 |
| Non-carriers | 68 (88%) | 70 (83%) |  |
| Carriers | 9 (12%) | 14 (17%) |  |
| Body mass index (kg/m<sup>2</sup>) | 28.24 ± 4.28 | 28.63 ± 3.57 | 0.5 |
| Mean arterial pressure (mmHg) | 97.87 ± 9.17 | 93.59 ± 9.95 | **0.005** |
| Anti-hypertensive treatment, n(%) | 31 (39%) | 66 (79%) | **<0.001** |
| 6-minute walk test (m) | 484.18 ± 74.04 | 568.79 ± 71.53 | **<0.001** |
| VO<sub>2</sub>peak (mL/kg/min) | - | 25.75 ± 5.05 | - |
| CBF (ml/100g/min) | 54.67 ± 11.09 | 54.87 ± 10.42 | >0.9 |
| Hippocampus CBF (ml/100g/min) | 41.44 ± 10.23 | 42.78 ± 8.40 | 0.4 |
| Posterior cingulate cortex CBF (ml/100g/min) | 58.90 ± 13.47 | 56.94 ± 11.29 | 0.3 |
| Anterior cingulate cortex CBF (ml/100g/min) | 50.22 ± 12.16 | 47.59 ± 9.74 | 0.13 |
| Precuneus CBF (ml/100g/min) | 48.31 ± 12.22 | 48.32 ± 10.47 | >0.9 |

Values are expressed as means ± standard deviations (SD), unless otherwise indicated. Baseline differences between groups were determined by t-student and chi squared tests for continuous and categorical variables, respectively, and significant differences are marked in bold. Abbreviations: CBF, cerebral blood flow; VO<sub>2</sub>peak, peak oxygen uptake.

### Pearson Correlations Between Cardiorespiratory Fitness, Cerebral Blood Flow and Covariates

In the AGUEDA sample, CRF indicator (6MWT) was not significantly correlated with the CBF variables studied (Figure 1), whereas in the Heart-Brain project, both CRF indicators analyzed (6MWT and VO_2_peak) were positively correlated with the CBF outcomes (Figure 2). Both cardiovascular risk factors studied (BMI and MAP) were negatively correlated with CBF in both samples (AGUEDA, BMI: *r*= -0.23, *P*= 0.039, MAP: *r*= -0.25, *P*= 0.025; Heart-Brain, BMI: *r*= -0.39, *P*<0.001, MAP: *r*= -0.23, *P*= 0.033). The negative correlation between age and CBF was stronger and significant in the AGUEDA sample, but not in the Heart-Brain sample (Figures 1 and 2).

**Figure 1.**
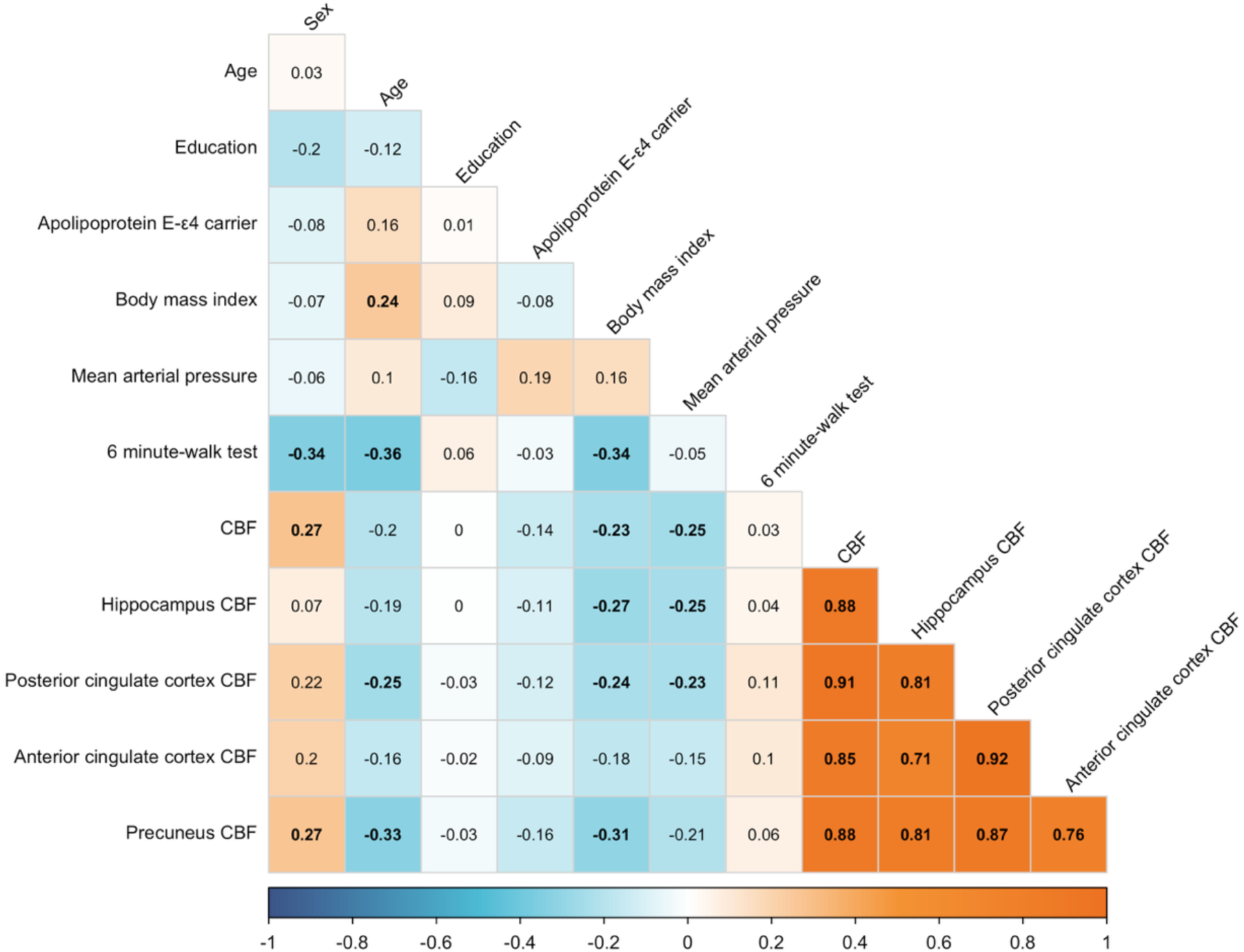
Matrix of Pearson correlation coefficients (r) between 6 minute-walk test, cerebral blood flow and covariates, for the AGUEDA sample. The colors of the boxes indicate the strength of the correlation, as shown in the explanatory scale on the bottom of the figure. Statistically significant correlations (p&0.05) were presented in bold. Sex was coded as 1=male and 2=female. Apolipoprotein E-ε4 carrier was coded as 1=allele carrier and 0=non-carrier. Abbreviations: CBF, cerebral blood flow.

**Figure 2.**
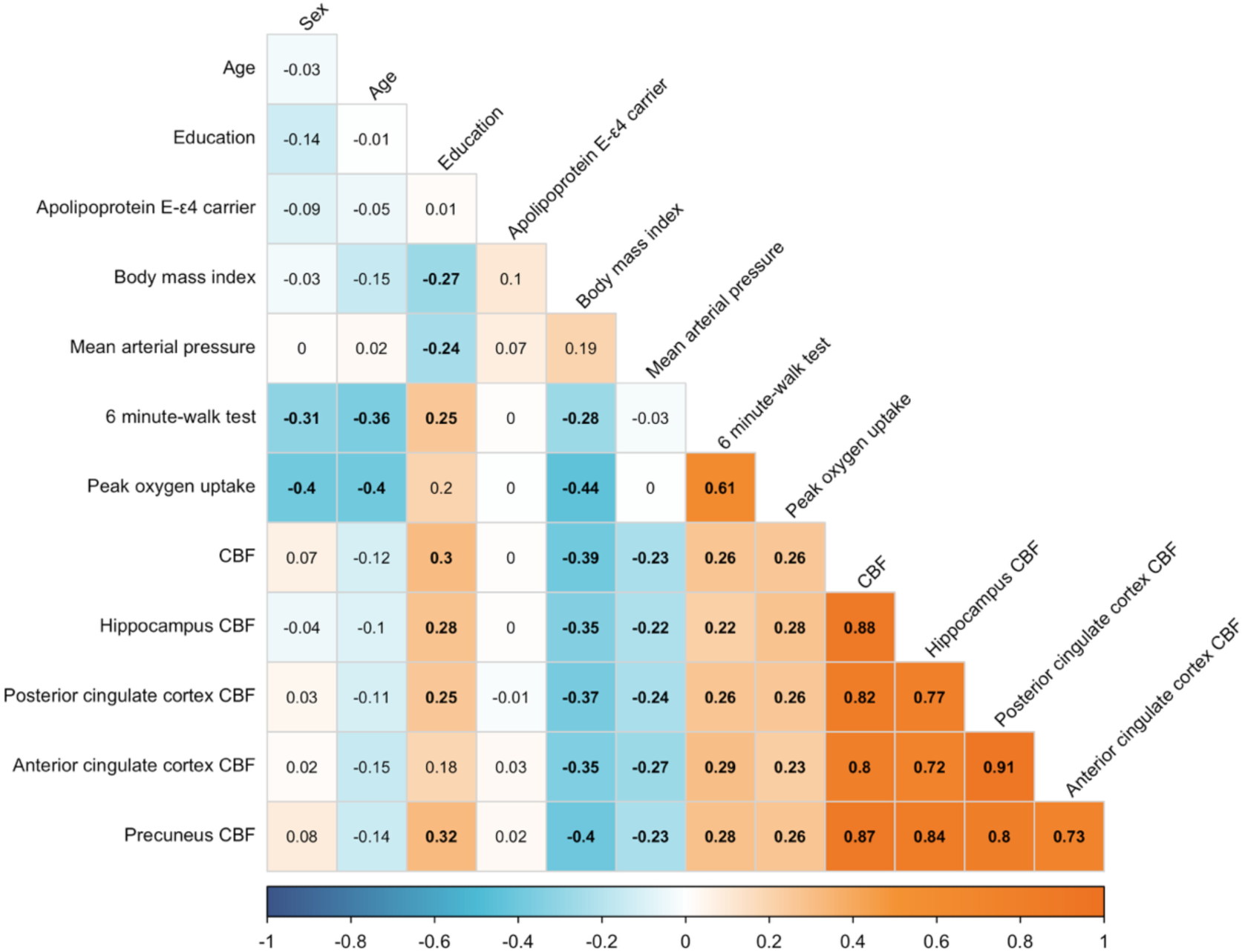
Ma trix of Pearson correlation coefficients (r) between 6 minute-walk test, cerebral blood flow and covariates, Heart-Brain sample. The colors of the boxes indicate the strength of the correlation, as shown in the explanatory scale on the bottom of the figure. Statistically significant correlations (p&0.05) were presented in bold. Sex was coded as 1=male and 2=female. Apolipoprotein E-ε4 carrier was coded as 1=allele carrier and 0=non-carrier. Abbreviations: CBF, cerebral blood flow.

### Association between 6-minute walk test performance and CBF

Figure 3 shows the associations between the 6MWT and CBF outcomes. Although positive trends were observed, when adjusting for age, sex, education, APOE4 carrier status and MAP, we found no significant association between 6MWT and CBF in the AGUEDA sample (standardized (Std.) *β*= 0.14, *95% CI* -0.10 to 0.38, *P* = 0.25), whereas a significant association was found in the Heart-Brain sample (*Std. β* = 0.26, *95% CI* 0.01 to 0.50, *P* = 0.04). Regional analyses did not yield significant associations between 6MWT and the brain regions studied for the AGUEDA sample (all *P* ≥0.05), whereas in the Heart-Brain sample significant associations were found for the posterior cingulate cortex (*Std. β* = 0.26, *95% CI* 0.01 to 0.50, *P* = 0.04), the anterior cingulate cortex (*Std. β* = 0.29, *95% CI* 0.05 to 0.54, *P* = 0.02) and the precuneus (*Std. β* = 0.28, *95% CI* 0.04 to 0.51, *P* = 0.02), but not for the hippocampus (*Std. β* = 0.16, *95% CI* -0.09 to 0.42, *P* = 0.20). The associations were markedly attenuated when adding BMI as covariate to the model.

**Figure 3.**
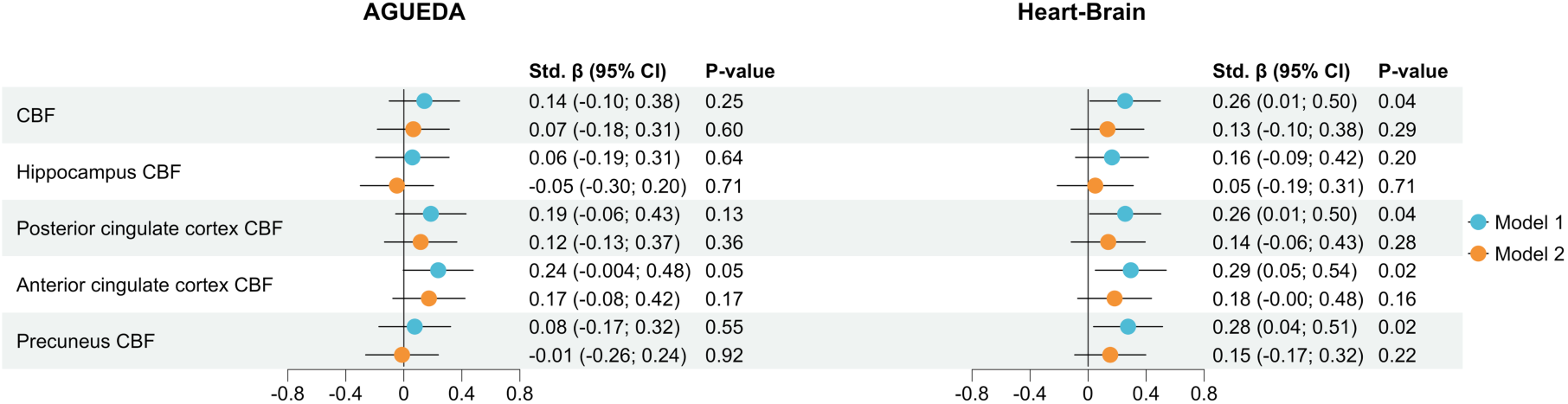
Association between six-minute walk test and cerebral blood flow. The results represent standardized beta coefficients from linear regression analyses, with 95% confidence intervals. Model 1 was adjusted by sex, age, education, apolipoprotein E4 carrier status and mean arterial pressure Model 2 was additionally adjusted by body mass index. Abbreviations: CBF, cerebral blood flow; CI, confidence intervals; Std, standardized.

#### Association between VO2peak and CBF

In the Heart-Brain sample, the VO_2_peak was positively associated with CBF, resulting significant for global CBF (*Std. β* = 0.33, 95% CI 0.07 to 0.58, P = 0.02; Figure 4A; Supplementary Table S2). Regional analyses showed VO_2_peak to be positively associated with CBF in the hippocampus (*Std. β* = 0.29, 95% CI 0.02 to 0.55, *P* = 0.03; Supplementary Table 2), the posterior cingulate cortex (*Std. β* = 0.31, 95% CI 0.05 to 0.57, *P* = 0.02; Supplementary Table S2) and the precuneus (*Std. β* = 0.32, *95% CI* 0.07 to 0.58, *P* = 0.01; Supplementary Table S2). These associations were markedly attenuated and became non-significant when adding BMI as a covariate to the model (Supplementary Table S2).

**Figure 4.**
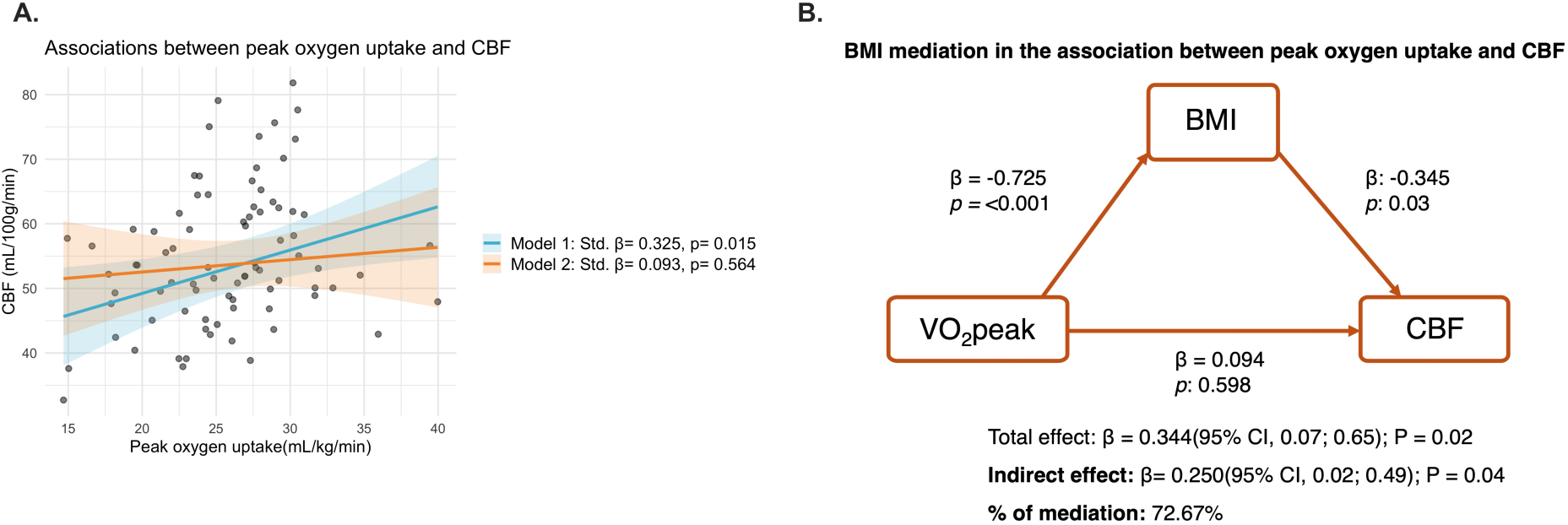
Association between peak oxygen uptake and cerebral blood flow in the Heart-Brain sample. **(A)** Dots show unstandardized values. Standardized beta coefficients and p values are shown in the legend. Model 1 was adjusted by sex, age, education, apolipoprotein E4 carrier status and mean arterial pressure. Model 2 was additionally adjusted by body mass index. **(B)** Mediation of the association between VO_2_peak and CBF by BMI for the Heart-Brain sample. Abbreviations: BMI, body mass index; CBF, cerebral blood flow; VO_2_peak, peak oxygen uptake.

#### Mediation effect of BMI in the association between CRF and CBF

In individuals with CAD (Heart-Brain sample), exploratory analysis showed that the association of VO_2_peak on CBF was largely mediated by BMI (Indirect effect: *β*=0.250, *95% CI* 0.02 to 0.486; percentage of mediation=72.67*%*; Figure 4B). When CRF was assessed by the 6MWT, the mediation effect of BMI was still significant individuals with CAD (Heart-Brain sample, indirect effect: *β*= 0.131, *95% CI*, 0.02 to 0.24; percentage of mediation= 47.12*%*). Despite not being significant in the population of cognitively normal older adults (AGUEDA sample, indirect effect: *β*= 0.077, *95% CI*, -0.01 to 0.17; percentage of mediation = 53.47*%*; Figure 5), the indirect effect and percentage of mediation was consistent and of similar effect size.

**Figure 5.**
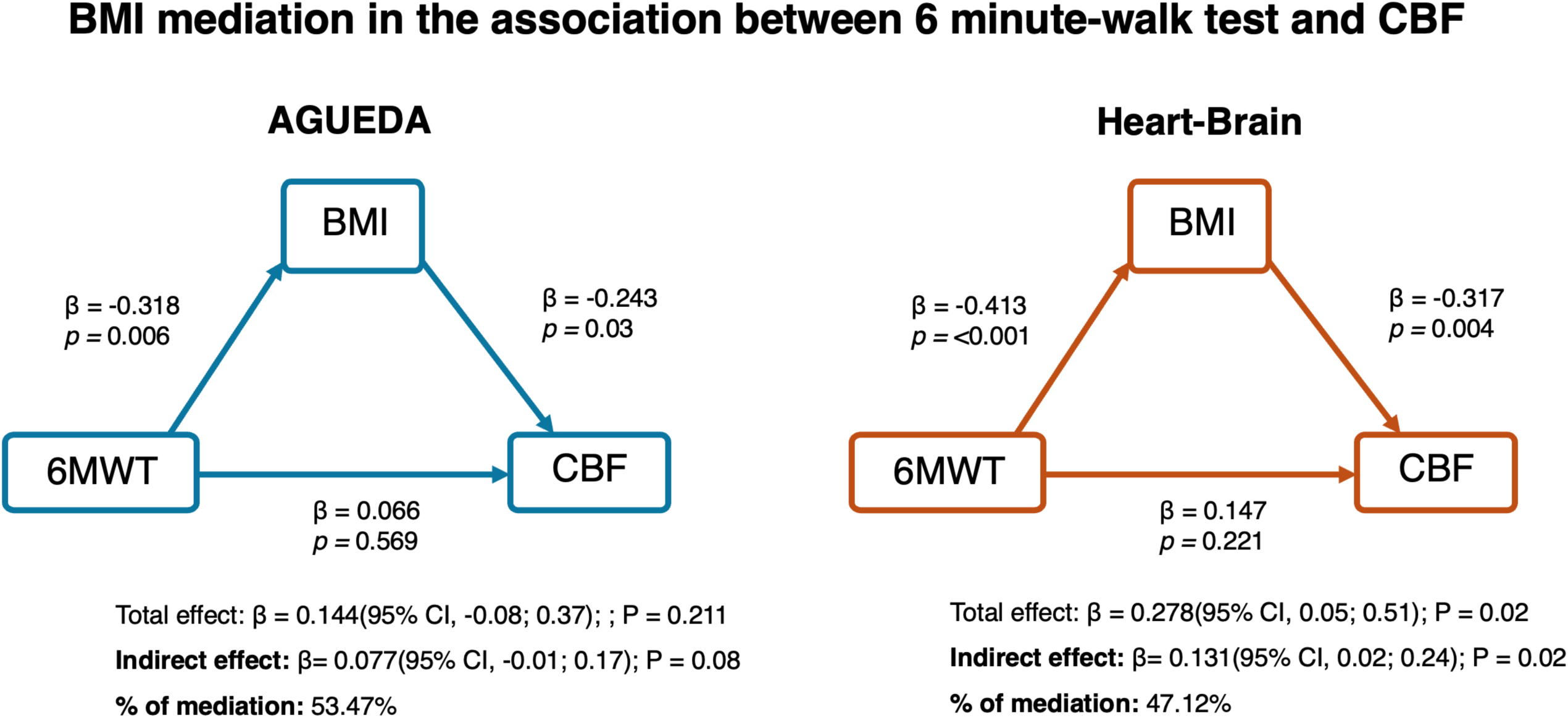
Mediation of the association between six-minute walk test and CBF by BMI in the AGUEDA and Heart-Brain samples. Abbreviations: BMI, body mass index; CBF, cerebral blood flow; 6MWT, 6-minute walk test

## Discussion

The present study aimed to examine the association between CRF and CBF in cognitively normal older adults and individuals with CAD. The main finding was that CRF was positively associated with CBF in individuals with CAD (Heart-Brain sample), but not in a sample of cognitively normal older adults (AGUEDA sample). In particular, performance on the 6MWT and VO₂peak were related to higher global and region-specific CBF in the Heart-Brain sample. Interestingly, these associations were dependent on BMI, which largely mediated the relationship between CRF and global CBF among individuals with CAD.

We found no significant association between 6MWT and CBF (global or regional) in the AGUEDA sample. Even though, we detected a positive trend for the anterior cingulate cortex, which has been previously shown to be responsive to exercise interventions.^74^ In line with our results, two previous studies in healthy older adults (aged between 60 and 89) reported either a negative association between VO₂peak and global CBF in unadjusted analyses^36^ or no significant association, after sex and age adjustment^36^ and after age, sex, blood pressure (systolic and diastolic), BMI, hand grip strength, and grey matter volume adjustments.^41^ Similarly, Krishnamurthy et al. found a positive but no significant relationship (no covariate adjustment), in cognitively healthy older adults.^40^ In contrast, one study revealed that VO_2_peak was positively associated with CBF, while accounting for variance explained by physical activity, age, gender, APOE4 status, Alzheimer’s Disease family history status, education and handedness, in 100 late-middle-aged adults (63.6±5.9 years).^38^

On the other hand, few studies have focused on or included individuals with CAD when testing the link between CRF and CBF. One study in 72 adults aged 50 or above, including 35 individuals with CAD, found a positive significant association for the whole sample, globally and in a wide range of regions. However, they found no interaction effect of the CAD condition, although the relationship was stronger for this subgroup.^75^ Just focusing on populations with CAD, only one study investigated the association between CRF and CBF, finding positive associations in the pre- and post-central gyrus, paracingulate, caudate, and hippocampal regions,^39^ in line with our results. We observed a positive and consistent association between CRF (both, 6MWT and VO_2_peak) and CBF in the Heart-Brain sample. At the regional level, greater 6MWT distance and higher VO₂peak were associated with higher CBF in the posterior cingulate cortex and the precuneus; the anterior cingulate cortex was associated only with 6MWT performance, and the hippocampus only with VO₂peak. These results indicate that fitter individuals with CAD have a better neurovascular health. The more consistent link between CRF and CBF in CAD may be attributed to the inherent poorer vascular condition. Previous age-matched studies showed that individuals with CAD had lower CBF than healthy controls.^17,20^ This is likely driven by atherosclerosis, endothelial dysfunction and reduced cardiac output^76–78^ -common conditions in CAD, although not exclusive to-, all of which are closely linked to VO_2_peak.^79–81^ Noteworthy, when BMI was added to the model results from both samples were drastically attenuated, not showing any significant relationship. We indeed detected that BMI could be acting as a potential mediator between VO_2_peak and CBF, although causal inferences cannot be drawn, as these analyses are based on one time point data. Obesity is a well-known cardiovascular risk factor,^82^ which has been also linked to accelerated brain shrinkage,^83^ more probable development of dementia^84^ and lower CBF.^24,41^ Hypotheses suggest that higher BMI may lead to increased secretion of pro-inflammatory cytokins from fat deposits, which can damage vascular beds, including those in the brain, and therefore impact the tissue’s health.^85^ As our model included MAP as a covariate, the effect of vessel damage was partially removed, indicating that other obesity driven pathways (e.g. oxidative stress, endoplasmic reticulum stress, sympathetic alterations, and metabolic dysfunction)^86^ rather than blood pressure, may play a critical role on CBF. Cross-sectional findings from Knight et al.^87^ showed that physical activity attenuated the association between BMI and CBF, highlighting the probable effect of lifestyle interventions on BMI reduction and CBF increments. Specifically, a previous 12-month physical activity and diet intervention demonstrated that CBF was positively associated with weight loss.^88^ Likewise, Espeland et al.^89^ found that CBF increments were linked to weight loss in type II diabetes individuals after an eight to 11 years physical activity and diet intervention, though results were not significant when comparing with the usual care group. Therefore, more interventions specifically targeting BMI reduction and CRF increments are in need to explore the potential effect of exercise, physical activity, diet and other behavioural interventions on CBF.

This study presents several key strengths. It conceptualizes the relationship between CRF, an important health marker, and CBF in two different populations at a high risk of cognitive decline and dementia, while investigating the potential mediating role of BMI. We have employed the 6MWT, which in our study showed consistent results as compared to the gold standard VO_2_peak, demonstrating that this test is a simple and informative approach for assessing CRF in large cohorts that aim to stablish cross-sectional and longitudinal associations with other health outcomes. Besides, for the CBF calculation we have employed multiple PLD ASL MRI which currently represents the most advanced, accurate, physiologically robust and repeatable non-invasive technique.^90^ Moreover, we revealed the importance of BMI on the relationship between CRF and CBF, warranting further longitudinal and experimental research focused on how weight reductions can elicit CBF increasement, especially in populations with cardiovascular or metabolic health conditions.

However, some limitations should also be acknowledged. First, although the 6MWT used in this study is well-suited for older or clinical populations due to its safety and feasibility, performance can be affected by pacing variability or participant’s motivation and it is less precise than gold-standard CRF measures such as VO₂peak, which was only determined for the Heart-Brain sample. Second, ASL MRI protocols across the literature present methodological variations [e.g. pseudo-continuous ASL (pCASL) versus pulsed ASL (PASL), single versus multiple post-labeling delays (PLDs), and differences in PLD timing or number of slices] which may affect cross-study comparability. Furthermore, the largest PLD -defined as the time between blood labelling and the image acquisition-in the pCASL sequence was 2 seconds, potentially not capturing the signal from regions where the blood takes longer to arrive. Finally, the multiple tests conducted may increase the likelihood of type I errors, and the cross-sectional nature of this study prevents causal interpretation. Longitudinal or intervention-based designs are needed to determine whether improving CRF leads to sustained increases in CBF and whether such changes are mediated by BMI variations.

In conclusion, we found CRF to be positively and consistently (using the 6MWT and VO₂peak as indicators) associated with CBF in individuals with CAD, but not in cognitively normal older adults, indicating that individuals with already impaired vascular function might benefit more from preserving CRF. Interestingly, the association of CRF with CBF was largely explained and mediated by BMI (+70%), indicating that fitter individuals with CAD have better brain perfusion, though this is seemingly attributed to their lower BMI. Nevertheless, these analyses were based on observational data, thus, causal inferences cannot be drawn. Future longitudinal and experimental studies are warranted to clarify the role of CRF and BMI in relation to CBF, the mechanisms involved and the implications for dementia risk prevention.

## Data availability

The data presented in this study is available upon reasonable request to the corresponding authors.

## Acknowledgements

The authors would like to thank the participants that took part in the two projects.

## Supplementary material

**Supplementary Table S1.** Results from the visual inspection and quality control on arterial spin labelling (ASL) images.

| Reason for ineligibility | AGUEDA | Heart-Brain |
| --- | --- | --- |
| Excessive motion resulting in spurious effects in difference map | 2 | 3 |
| Differences in perfusion between hemispheres | 1 | 1 |
| Brain territories without perfusion signal | 2 | 2 |
| No perfusion signal due to wrong placing of the ASL imaging slab | 2 | 3 |
| Grey matter CBF coefficient of variance | 4 | 3 |
| **Total images discarded** | **11** | **12** |

Data represent the number of ASL images discarded by project and reason for ineligibility
Abbreviations: ASL, arterial spin labelling; CBF, cerebral blood flow.

**Supplementary Table S2.** Association between VO2peak and cerebral blood flow.

|  | Model 1 |  | Model 2 |  |
| --- | --- | --- | --- | --- |
| | Std. $\beta$ (95% CI) | P | Std. $\beta$ (95% CI) | P |
| CBF | 0.33 (0.07; 0.58) | 0.01 | 0.09 (-0.23; 0.41) | 0.56 |
| Hippocampus CBF | 0.29 (0.02; 0.55) | 0.03 | 0.10 (-0.24; 0.43) | 0.57 |
| Posterior cingulate cortex CBF | 0.31 (0.05; 0.57) | 0.02 | 0.10 (-0.23; 0.42) | 0.56 |
| Anterior cingulate cortex CBF | 0.25 (-0.02; 0.51) | 0.07 | 0.00 (-0.33; 0.33) | 0.99 |
| Precuneus CBF | 0.32 (0.07; 0.58) | 0.01 | 0.08 (-0.23; 0.40) | 0.60 |

The results represent standardized beta coefficients from linear regression analyses, with 95% confidence intervals. Model 1 was adjusted by sex, age, education, apolipoprotein E-ε4 carrier status and mean arterial pressure Model 2 was additionally adjusted for body mass index.
Abbreviations: CBF, cerebral blood flow; CI, confidence intervals; Std, standardized.

